# Modeling Exit Strategies from COVID-19 Lockdown with a Focus on Antibody Tests

**DOI:** 10.1101/2020.04.14.20063750

**Authors:** Reinhard German, Anatoli Djanatliev, Lisa Maile, Peter Bazan, Holger Hackstein

## Abstract

This paper presents two epidemiological models that have been developed in order to study the disease dynamics of the COVID-19 pandemic and exit strategies from the lockdown which has been imposed on many countries world-wide. A strategy is needed such that both the health system is not overloaded letting people die in an uncontrolled way and also such that the majority of people can get back their social contacts as soon as possible. We investigate the potential effects of a combination of measures such as continuation of hygienic constraints after leaving lockdown, isolation of infectious persons, repeated and adaptive short-term contact reductions and also large-scale use of antibody tests in order to know who can be assumed to be immune and participate at public life without constraints. We apply two commonly used modeling approaches: extended SEIR models formulated both as System Dynamics and Agent-Based Simulation, in order to get insight into the disease dynamics of a complete country like Germany and also into more detailed behavior of smaller regions. We confirm the findings of other models that without intervention the consequences of the pandemic can be catastrophic and we extend such findings with effective strategies to overcome the challenge. Based on the modeling assumptions it can be expected that repeated short-term contact reductions will be necessary in the next years to avoid overload of the health system and that on the other side herd immunity can be achieved and antibody tests are an effective way to mitigate the contact reductions for many.

## 1 Introduction

In December 2019 an outbreak of pneumonia in Wuhan, Hubei, China has been detected. It has been caused by a new coronavirus, named now severe acute respiratory syndrome coronavirus 2 (SARS-CoV-2) by the World Health Organization (WHO). The disease, now referred to as coronavirus disease 2019 (COVID-19) by WHO, ran up to a world-wide pandemic: on April 13, 2020, the well recognized Johns Hopkins University dashboard^1^ [1] reported ca. 1.8M confirmed infections, 115K deaths, and 430K recovered. A major problem is the sudden overload of the health system with people needing hospitalization, intensive care units (ICU), or ventilation, at the same time. If this happens, people can not access the help they need and die in an uncontrolled way. In face of that, many countries world-wide have taken preventive isolation as well as social distancing measures, mainly in order to avoid such an overload and to flatten and shift the peak stress for the health system, also in order to be able to prepare for the wave of demand. Another major concern is that personnel in the health system and in other system-relevant sectors must be protected against infections.

However, this results in a lockdown of public life and causes many other problems, besides the significant restriction of civil rights it is a major challenge for the economy, income of people, and other negative effects. Therefore, it is obvious that such a lockdown must be restricted to periods as short as possible and it is a question of utmost relevance how it is possible to exit from this lockdown such that

- the peak of health care demand is mitigated so it can be handled by health care resources and
- contact reduction measures can be relaxed and people can as soon as possible return to normal life.

Undoubtedly, a combination of measures will be required to return safely and in steps to normal life, such as quarantine of infectious persons, continued control of fatalities and potential repetition of contact reduction measures if they get too high again, risk-group-adapted partial relaxation of contact reduction, and continued hygienic constraints. App-based digital contact tracing has also been suggested for epidemic control [2] but is controversial due to privacy issues. Of specific importance to guide this process are antibody tests in order to know which persons can be assumed to be immune and, thus, can return to normal life. Such tests are under development but currently no significant capacities do exist. It is therefore an important question to find

- requirements for such tests in terms of test capacity, as well as adequate sensitivity and specificity values
- strategies how these tests can be applied in order to be effective.

Additionally, it must be taken into account that the disease is asymptomatic or shows just mild symptoms in the majority of cases and can lead to immunity for many (herd immunity). If the lockdown is left too early or in an uncontrolled way, infections can rebound and successive interventions can become necessary. This should be kept to a minimum.

Currently, an increasing variety of SARS-COV-2 antibody tests is available. From a medical point of view, seroconversion, i.e., positivity of IgM/IgG antibodies can be detected as early as 7-14 days after symptomatic COVID-19 infection [3]. However, since many patients can still shed oral infectious virus up to 22 days after onset of symptoms or after first positive COVID-19 PCR test result [3], sole positivity of combined IgM/IgG is not a good indicator for non-infectious individuals and herd immunity. In this respect, isolated detection of virus-specific IgG should be preferred, because it has a delayed kinetic in comparison to IgM. Furthermore, with respect to disease modeling and contagiousness a safety buffer of 14 days added on the date of IgG positivity may be reasonable in order to define a reliable time-point when a person should be considered recovered and non-infectious. Another important variable of antibody tests is their specificity and sensitivity. For the reliable calculation of herd immunity a high specificity of an antibody test is critical and should be >99%. Also, an extremely high sensitivity of the antibody test might be desirable and nice to have. However, it is not of critical importance, as it can be assumed that most people with a functioning herd immunity also have sufficiently high antibody levels.

In order to find strategies to defeat the pandemic, epidemiological models are used to support decision making. In general, there are two main modeling approaches: an aggregate view based on a system of differential equations, also known as System Dynamics (SD) and an individual-based simulation, also known as Agent-Based Simulation (ABS) [4]. SD models can describe dynamics on an abstract level, people in certain states are represented by their number and for the solution just an ordinary system of differential equations needs to be solved. Standard solvers are available, the size of the system is normally quite small (up to a few ten equations) and a fast, immediate response is possible. In ABS, each individual is modeled explicitly allowing for stochastic and more detailed behavior, underlying is however discrete-event simulation causing higher computational costs (note that also repetitions of the simulations are needed in order to get statistically reliable results). Depending on the goals a suitable modeling approach can be selected. It is also possible to combine both, as an example of hybrid simulation, which has already been applied successfully in healthcare simulations [5]. It should be noted that an SD model corresponds to an ABS where all agents are represented by a Markov chain with the same states as the SD model and the number of agents is taken to infinity [4]. Therefore, all timing in SD is implicitly exponentially distributed. By splitting the SD variables it is also possible to represent phase type distributions such as the Erlang distribution which is less variant [6]. This allows for more realism in SD models. Most models extend well-known SEIR models [7, 8] with the states susceptible, exposed (infected but not yet infectious), infectious, and recovered. In SD models each equation describes the change of the number of people in theses states and in ABS each agent has these internal states.

A number of simulations with both approaches for the dynamics of COVID-19 have been published recently and are discussed in Sec. 2. These models describe the dynamics of the disease and the effects of certain interventions. For this purpose efforts have been undertaken to get important model parameters such as basic reproduction number, incubation period, case fatality risk, others cannot be accessed and must be estimated, e.g., percentage of contact rate reductions when interventions are implemented or the percentage of isolated cases. It is however clear that at this stage of the pandemic there are many uncertainties about these parameters, including the manifestation index, fatality rates as well as age- and risk-stratified numbers. In this paper we use those models as a reference in order to adequately model the disease dynamics, adapt them to current data and then add further aspects for investigating exit strategies based on a set of measures.

The contribution of this paper is to investigate for a country like Germany combinations of measures in order to get an effective exit strategy and to meet the important goals mentioned above. We will thus investigate the following scenarios: disease dynamics if no lockdown would have been imposed, an exit of the lockdown without successive measures, hygienic constraints imposed after lockdown, and repetitive and adaptive but short-term social contact restrictions. Isolation of infectious people, seasonality of contagiousness, and immunity of a large fraction of the population is also considered. Antibody tests are used in order to let return people with assumed immunity to public life without contact reductions. Since such tests are a scarce resource it will not be possible to test all people systematically. Therefore it will be necessary to develop strategies such that a positive effect can be achieved. One approach is to give people from risk groups or from system relevant sectors preference, such as health personnel. A second approach is to follow infection chains and identify people who might have been infected but were asymptomatic (digital contact tracing could help here). If that is possible, these people will be also tested with preference leading to a higher fraction of people able to reenter public life. We will first examine these strategies with an extended SD model in order to get main hints. We will then show an ABS model for a prototypical region with different classes of people. The ABS model will be used in followup work to check whether properties of testing are sufficient or must be improved. In the SD model all effects are aggregated in rates for the complete population, whereas in the ABS individual contacts between people in families, at work, in hospitals, and in leisure are modeled. Thus the ABS allows to draw conclusions also for the availability of personnel in hospitals and in companies. This also allows for planning over the time during and after lockdown. Both models have been realized with the simulation framework AnyLogic^2^.

The rest of the paper is organized as follows. In Sec. 3 we discuss available data sources, uncertainties within them and extract our assumptions which we use in both the SD and ABS models. Sec. 4 presents the SD and Sec. 5 the ABS model. Results are given Sec. 6, conclusions and further work are discussed afterwards.

## 2 Related work

A number of simulations for the dynamics of COVID-19 have been published recently. SD models for the dynamics of COVID-19 and the effects of possible interventions are for instance [9], which is accessible online^3^, the model of Robert Koch Institute for Germany [10] which we consider as a reference model for the study presented here. In [11] an ABS model is presented for studying the dynamics of COVID-19 and possible mitigation and suppression measures in GB and US, a model representing each inhabitant of Austria^4^ is based on [12].

A multiple-input deep convolutional neural network model is used in [13] to predict the number of confirmed cases in China with respect to the number of cases from the past five days. However, no measures such as contact restrictions or quarantine can be taken into account, but these have a significant impact on the spread of the virus and can lead to a subexponential growth in the number of cases. Using China as an example, this influence is examined in [14] with an extended SIR model and in [15] with an extended SEIR model. The agent-based simulation model [16] examines the influence of interventions on the spread of the virus in Singapore. To predict the local and nationwide spread of the virus, [17] combines a SEIR model based on differential equations with a metapopulation model based on traffic flows to model intercity mobility. The influence of traffic restrictions on an international level is examined in [18] with a combined individual-based subpopulation and a flow-based metapopulation model.

In conjunction with testing, measures to reduce the probability of transmission can be improved. In [19] the influence of the test coverage with regard to the deceased on the contact rate with an extended SEIR model is examined. According to the calculations in [20], carrying out mass tests can significantly reduce the economic costs of mitigating the COVID-19 pandemic. For this purpose, the SEIR model is expanded to differentiate between recognized and unrecognized infected people. The influence of such tests can also be tried out using online available SD models^5,6^. A particular limiting factor is the availability of test kits. The pool testing strategy for asymptomatic or mild cases presented in [21] can reduce test costs and improve the identification of low-risk individuals under the condition, that most tests are negative.

The models without tests for infected persons partially combine the contact reduction to reduce the infection rate with measures such as isolation or school closure. However, no strategies for adapting these measures are modeled. The models which consider tests use this extension to continuously adjust the contact rate. Additionally, the study [20] also switches between phases with a low and phases with a high test rate. All models have in common that they do not take into account tests for antibodies.

## 3 Data sources, assumptions, and extended SEIR model

There is a variety of information available about epidemiology of COVID-19, by WHO and in many preprints and accelerated publications, most of them report insights from Hubai, China. We use the investigations of the Robert Koch Institute (RKI) which is Germany’s public health institute. It continuously monitors the literature and gives a characterization^7^ of facts and numbers for which some evidence is available, last updated on April 10, 2020. It is clear that at this stage there is much uncertainty in many of those values, but we extract the main findings which are relevant for modeling. For some required figures RKI does not give characterizations, in these cases we resort to the assumptions of [10], which are also based on reflections from experienced epidemiologists. Both information sources are (unfortunately) in German. The presented values are default values which can easily be changed, as will be done in later experiments.

The *manifestation index* of showing symptoms when infected is estimated to be within 69% and 86%. However, this might be larger, some estimations even assume a factor of unknown cases of up to 20. *Severity* is characterized as mild or moderate (without or with light pneumonia, 80%), severe (needs hospitalization without ICU, 14%), and critical (needs ICU 6%). The *basic reproduction number R*_0_ is estimated to lie between 2.4 and 3.3, WHO estimates for China a range between 2 and 2.5 (without interventions). We assume a value of 3 to reflect the recorded cases in Germany. As in [10] we also allow for seasonal changes according to a sine function. For the *incubation period* (from infection to sickness) on the average 5 to 6 days can be assumed (range 1 - 14). For the *latency period* (from infection to being infectious) no results are given, we thus assume 3 days. The *infection period* can be assumed to start 2.5 days before onset of symptoms, the duration is unclear. Since the severity and duration of cases differ we calculate the infection period weighted by the number of cases (both symptomatic and asymptomatic), resulting in an average of 12.5 days including the prodromal period. *Symptomatic period* in mild or moderate cases is assumed to be 9 days according to [10], we take the same time for asymptomatic cases. For the time from onset of symptoms to hospitalization an average of 4 is reported (with an interquartile range IQR of 2 - 7) and from hospitalization to ICU on the average 1 day (IQR 0 - 3). For the *hospitalization time* of severe cases and for the time in ICU we assume 14 and 10 days respectively as in [10]. *Case fatality rates* (CFRs) are counted in different ways and are uncertain, in [10] it is assumed that only ICU patients die with a percentage of 50%, in the RKI characterization also CFRs of 0.1% (mild) 8.1% (severe), 22% (critical) are suggested, we count moderate cases to be included into the mild ones. After the disease people have antibodies and are assumed to be immune. Even though the persistence is unclear, experience from other corona viruses suggest ca. 3 years, we assume here persistent immunity.

Based on this, a state chart for the progression of the disease from the view of a single person can be defined, it is shown in Fig. 1 and represents an extended SEIR model. Persons are initially *susceptible*, then *exposed* after infection, after the latency period a person gets *infectious* and is *recovered* or *dead* afterwards. Persons in recovered are assumed to be immune and cannot be infected again. The state infectious has substates: first there is the state *prodromal* in which no or only mild symptoms are shown, afterwards there is a split between *asymptomatic* and *symptomatic* cases. The symptomatic ones are further split into *mild, severe*, and *critical*, the last two of them are again subdivided into sequential states *at home, hospitalized*, and *ICU*. We also divide the recovered state into the substates *was_asymptomatic* and *was_symptomatic*, a separation which we need for the antibody tests. At each time instance the state chart is in exactly one state, for every (sub-)state without substates either a sojourn time distribution (e.g., latency period in *exposed*) or an event (e.g., infection by another person in *susceptible*) for leaving the state must be defined. In case of more than one outgoing arrow of a state, switching probabilities are required. For instance, persons leaving the substates of state *infectious* have different probabilities to get either back to susceptible (if the person is not immune afterwards, by default this probability is set to zero) or to *recovered* or *dead*. All necessary information for such sojourn times and switching probabilities can be obtained from the data mentioned in the previous paragraph.

**Figure 1:**
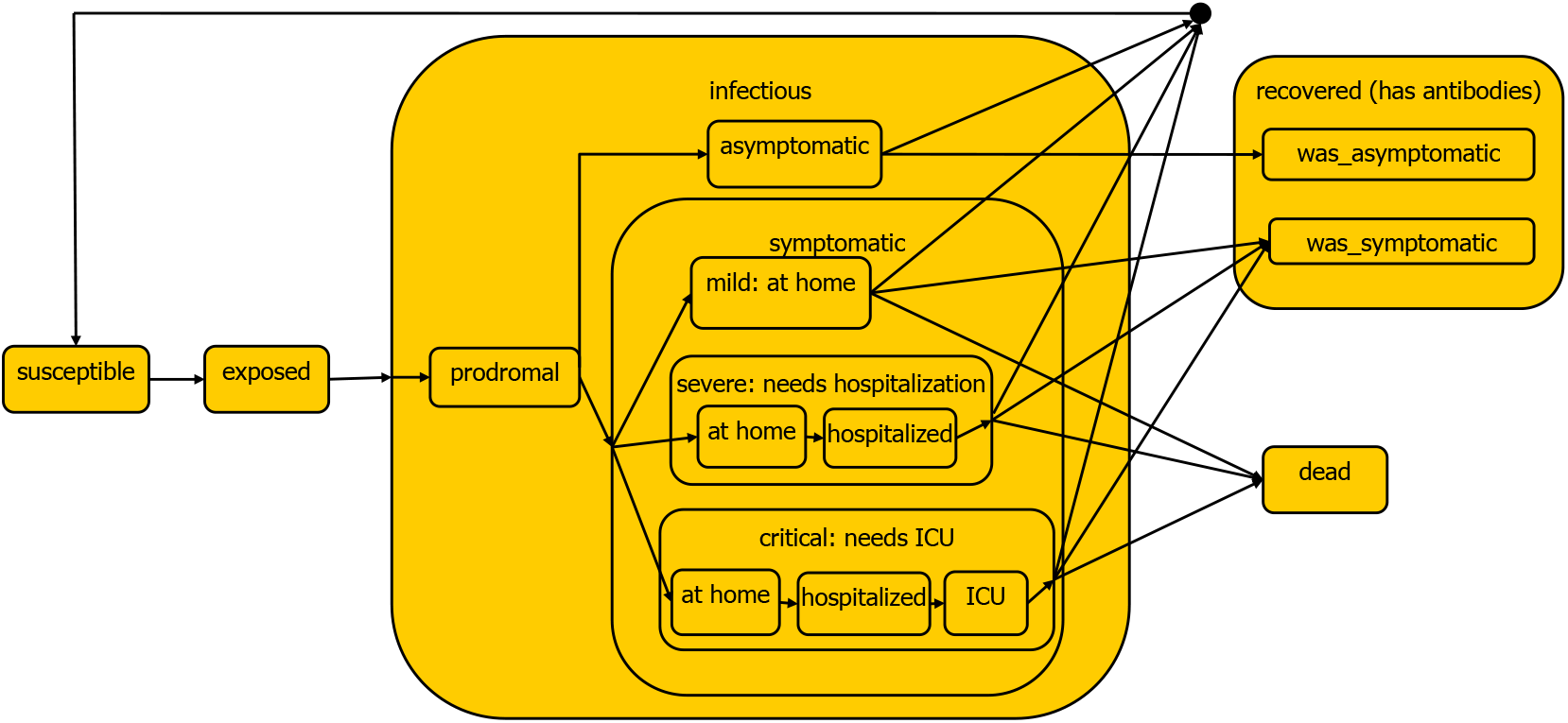
Extended SEIR model for the progression of the COVID-19 disease from the perspective of a single person

In order to model also aspects like governmental interventions and testing, further attributes are required. Every person has a *social status*: *normal, reduced*, and *isolated*; initial social status is *normal*. Considered governmental interventions are:

- *Reduction*: by assigning *reduced* to many persons, their contact rate is reduced; this models the actual lockdown. Without differentiation between population groups, a global percentage of contact reduction can be assumed, with 50% as a default value. With differentiation, we can assign different contact reduction percentages to, e.g., health personnel, employees in system-relevant sectors, and others. This affects the rate of infections significantly. Reduction is applied for persons in every state besides those in which a person is *isolated* or has regained the social status *normal* because of testing (see below). It is possible to define one reduction with its starting time and its duration. Additionally, it is possible to start and stop threshold-based reductions: when the CFR gets above or below a definable threshold, respectively.
- *Hygienic constraints*: when *reduction* is taken back, continued hygienic measures are likely to be implemented, e.g., mandatory respiratory masks in public life. In the model this causes a contact reduction with a lower percentage than in the previous case.
- *Isolation*: Persons in substates of *symptomatic* can be *isolated* by moving them in specialized isolation wards, persons in *ICU* are *isolated* anyway. For *isolated* persons the contact rate is set to zero. Persons in state *symptomatic* staying at home or in hospital get reduced contact rates which can be adjusted (default values are zero in both cases). This intervention is assumed to be permanent.

Testing both for COVID-19 and for antibodies (ABs) will be important for an exit of the lockdown. For the time being we restrict ourselves to AB tests. Every person can be *tested* (in principle possible in all states, but testing would not be meaningful in state *symptomatic*), with either *positive* or *negative* outcome. We assume that persons with ABs are immune and will keep that property indefinitely (could be relaxed later). Therefore, *positive* ones, regardless in which state besides *symptomatic*, can change their social status from *reduced* to *normal*. We consider the sensitivity (by default 95%) of the test, leading to probabilities for true positives and false negatives (*negative* ones have ABs but would stay in *reduced* although not necessary). We assume also a high specifity and ignore false positives (*positive* ones without ABs would get *normal* but can still be infected), because this number is expected to be small. Test capacity is an important restriction and will be adjusted in the experiments, test duration is assumed to be one day.

A hypothetical unlimited test capacity constitutes a benchmark to investigate the maximum benefit from AB tests. Since test capacity is however limited, different strategies to allocate tests are conceivable. A first strategy could be to give preference to persons in state *was_symptomatic* and allocate possible leftover capacity to all others randomly. A drawback of this strategy is that most test capacity would be used for symptomatic recovered, who will probably have ABs. An effective strategy would be to use tests mainly for persons in state *was_symptomatic*, because this can be many and they can change their social status from *reduced* to *normal*. Given it is possible to identify them, they should get a preference. Such an identification is to a certain degree possible by following infection chains and could be even more accurate by digital tracing. In order to represent such strategies in a flexible way, we will use percental weights for persons in the different states. E.g., a complete random strategy would allocate the same weight to all states, whereas a strategy giving preference to persons in *was_symptomatic*, leaving no capacity for persons in *was_symptomatic*, and distributing smaller capacities to others without symptoms could be: *was_asymptomatic* → 40%, *was_symptomatic* → 0%, *asymptomatic* → 20%, *prodromal* → 15%, *exposed* → 15%, *susceptible* → 10%. The probability of a person to get tested is proportional to the number of persons in the same state and the weight.

## 4 System dynamics (SD) model

Based on the extended SEIR model of Sec. 3 an SD model is derived and shown in Fig. 2. SD models consist of stocks (continuous variables representing the number of persons in this state) and flows between them (associated with rates between the stocks). It is possible to derive flow rates from the sojourn times and switching probabilities presented in Sec. 3. Each colored rectangle corresponds to a stock with a name derived from the state chart in Fig. 1, each arrow corresponds to a flow, shorter names are also given for the underlying mathematical equations. The equations define the model precisely and are given with all parameters in Appendix A.

**Figure 2:**
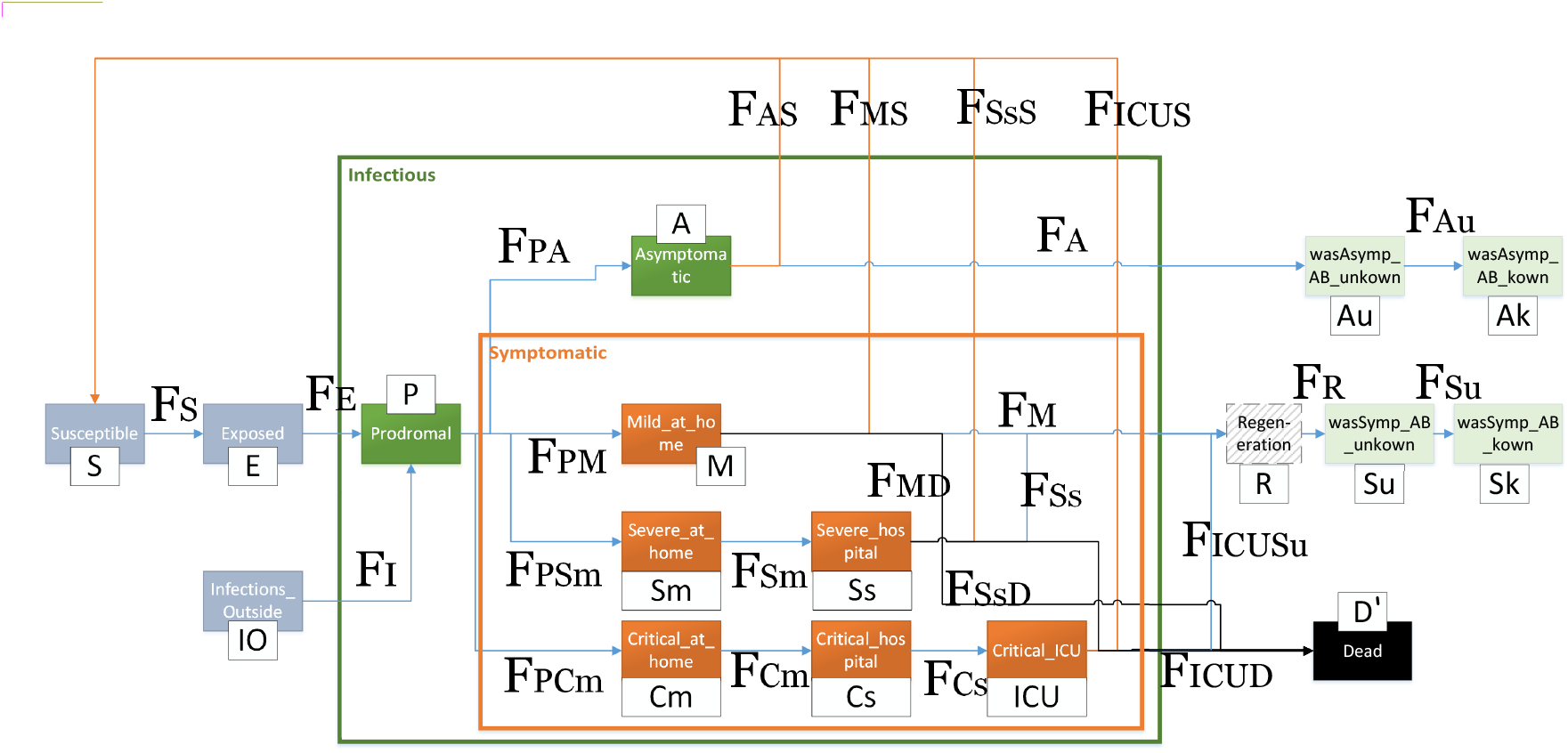
SD model for the COVID-19 disease

Note that the two states *was_asymptomatic* and *was_symptomatic* are split into 4 stocks: *wasAsymp_AB_unknown, wasAsymp_AB_known, wasSymp_AB_unknown*, and *wasSymp_AB_known*, in order to separate between cases where recovered persons have ABs but this is not known vs. cases in which it is known because of AB tests. In case of asymptomatic and recovered persons this is completely unknown and in case of symptomatic and recovered persons it can be supposed that they have ABs but without certainty. There is also an additional stock to represent infections coming from outside. The stock *Regeneration* is only implemented to account for the fact that people are considered as recovered with a delay. It is used since patients after being sick cannot directly participate in public life and its duration is set to 5 days to reflect values in Germany. However, it can as well be omitted, since it only delays statistics for recovered people. Flow *F*_*Au*_ corresponds to true positive AB tests of recovered with ABs who were asymptomatic whereas flow *F*_*Su*_ corresponds to those who were symptomatic. For the corresponding flow rates factors need to be computed consisting of the sensitivity, the proportion of the outgoing stock, and the weight mentioned in Sec. 3. These factors need to be normalized and multiplied with the available test rate, for details please check the Appendix.

There are several possible paths to extend the SD model. First, PCR tests could also be integrated. Second, to make timing more realistic, the implicit exponential distribution could be replaced by the Erlang distribution by splitting stocks into sequences. Third, to allow for sub-populations inside the model, the stocks could also be split. All extensions would make modeling more complicated but would not be challenging for the numerical solution.

## 5 Agent-Based Simulation (ABS) Model

This modeling approach allows to implement a more realistic simulation. In this case each person is represented by an agent which has individual attributes and follows one or more behavior models. In our ABS model agents are combined in different groups which are related to the locations where they can meet and infect each other considering group-related probabilities. Those are home/family, leisure (including activities like shopping, sports, etc.), being at hospital, and work. Depending on the current contact reduction level they can change their states. In case of isolation the agent remains at the quarantine location without changing its state. In order to evaluate a profession-based exit from the lockdown, different types of workers are considered, namely, system-relevant, not system-relevant, hospital staff, and others. In particular, the latter represent the non-working population including children and retired persons. Each worker is linked to a different sized company with other workers in order to avoid direct infections between the whole working population. The total number of companies is an input parameter that allows to generate large and small companies. Similarly, different sized households are built and persons are assigned to one certain home group. Healthcare workers can meet their colleagues, but also patients who are potentially infected. The infection probability between patients and healthcare personnel can be pre-configured separately from the infection between colleagues.

A further state chart represents the disease states of agents. Beginning by susceptibles, the statechart shows a behavior which is very close to the conceptual SEIR model of Sec. 3. By configuring the daily capacity for antibody tests, they can be applied following one of the defined strategies in Sec. 3. As the ABS model differentiates between working groups and other individual attributes, an even more precise testing strategy can be evaluated. For example, hospital staff and system-relevant workers can be tested with a higher priority. Input configuration allows to preset a sensitivity and a specificity value for the test. For a more precise analysis, it is even possible to consider different error probabilities for agents that are affected and have not been recovered yet. Fig. 3 shows a screenshot of the model with 20.000 agents, the agents are shown as icons in their environments, colors represent their states. The model can deal with several ten thousand agents easily and is ready to study effects in a region, but differentiated for the agent groups. It is for instance possible to derive figures of health personnel with ABs. It will also easily be possible to include other important aspects into the ABS model: age-dependent severity and CFRs, controlled change of social status (from *reduced* to *normal*) depending on vulnerability (e.g., elderly and other risk groups), viral load at infections (can influence severity), etc. Resource-based analysis can show, if enough beds, hospital staff, or system-relevant workers are available when the pandemic gets more severe. It would also be interesting to model regional clusters with ABS and connect them via continuous flows or to provide an SD environment with the general disease dynamics for the ABS model of a region, similar as in [5].

**Figure 3:**
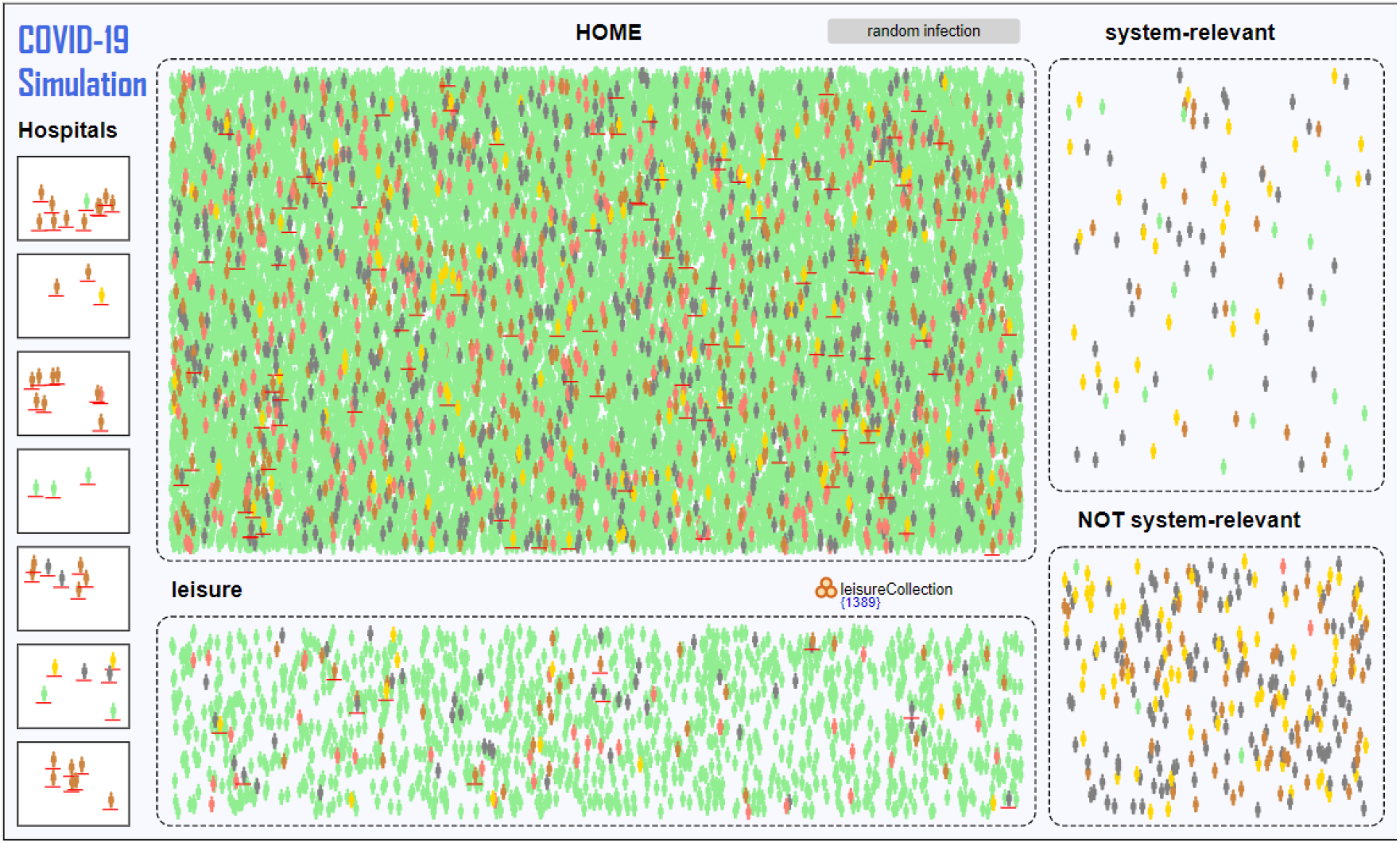
Screenshot of the ABS model for the COVID-19 disease

## 6 Results

We now present the results obtained with the SD model. We calibrated the model to reflect the real progress of the disease in Germany while staying as close as possible to the RKI values. Fig. 4 shows the model results with this calibration, the parameters are provided in the Appendix. The left upper curve show a *contact index* which we have defined in order to illustrate the overall effect of the interventions on the social contact abilities of people. It gives the contact abilities averaged over all people who are not symptomatic: if there would be no intervention, all people would have full contact abilities and the index would be equal to one. Maximum values of critical figures over the simulated time period such as the number of hospitalized or the number of patients needing ICU are shown in the right upper bar chart. In the middle row the progress of relevant figures such as number of symptomatic, other infected and exposed persons is shown on the left and daily deaths, ICU, and hospitalization needs on the right. The lower row shows on the left cumulated numbers of deaths and people who had been symptomatic. On the right, the grow of the number of persons is shown for which immunity can be assumed (if this number reaches ca. 60%, herd immunity can be assumed). The curve is split into known and unknown ones. The unknown ones are constituted in principle by those who had been symptomatic and are recovered or an AB test has been performed for them. In Fig. 4 no such AB tests are considered, but below we will investigate effects of AB tests.

**Figure 4:**
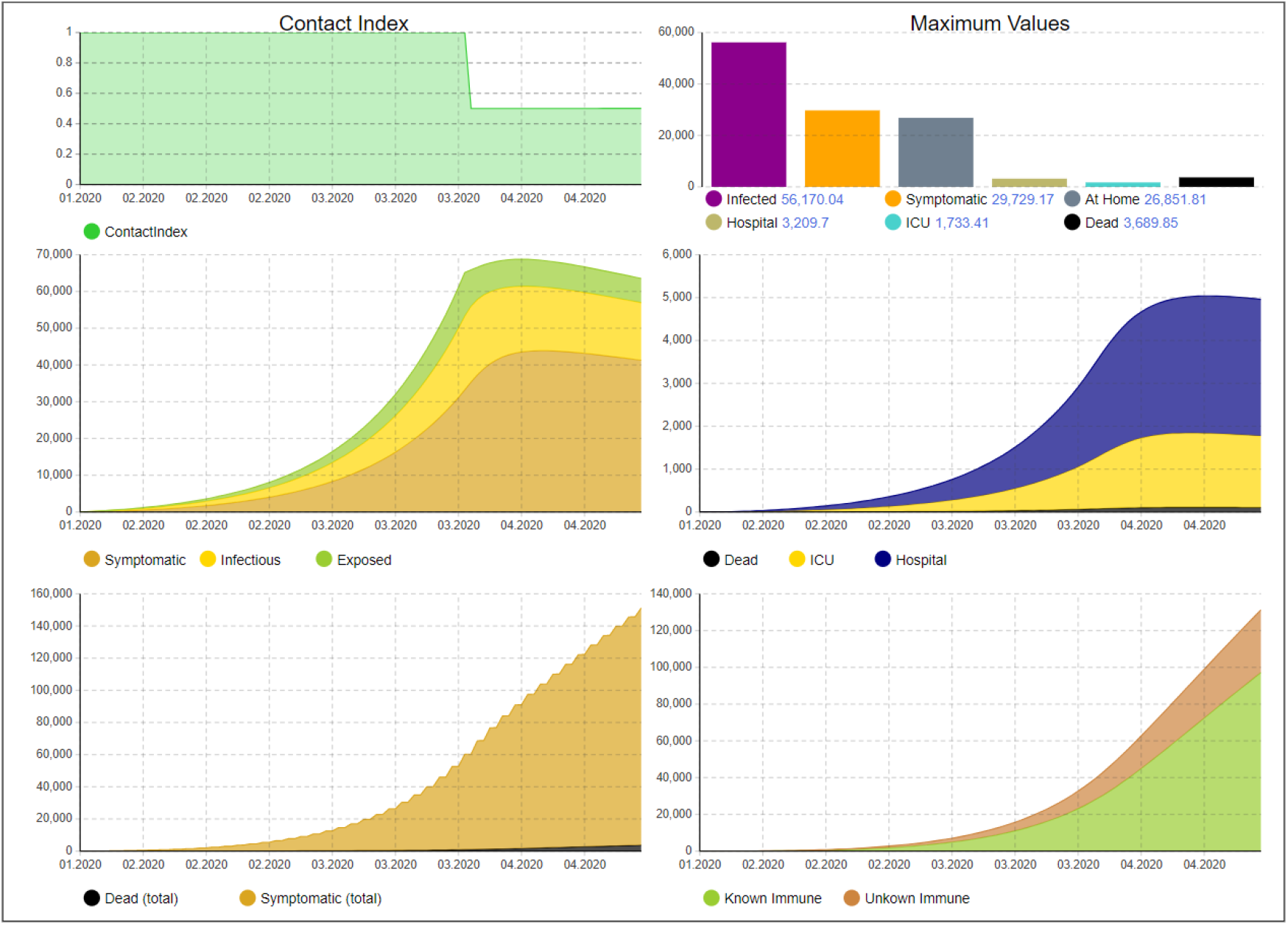
Disease dynamics until April 19

Compared to RKI, we assume a higher ICU capacity due to the report of hospital utilization (ca. 27K ICU places)^8^. The reproduction number is set to 3, meaning that on average, one infected person can spread the virus to three other persons. However, if the infected persons get symptomatic and stay at home, their contact rate is halved such that they only can infect half the number of persons. If they are isolated in a hospital, we assume that they infect on average one additional person, for example in their family, before they are isolated. We also slightly increased the number of people that require treatment in hospitals and ICUs to model the number of confirmed cases in Germany. The evolution of the virus in Germany has three distinctive time instants: 1) it started on January 22, 2) the majority of interventions have been introduced after March 22, and 3) the latest available data time when this paper has been written. We reflect the real number of infections and deaths according to the statistics provided by Johns Hopkins University^9^. The model tries to match the situation at all three times instants as close as possible. However, it is clear that we can only model an idealized behavior. We match the cumulative number of infections, deaths, and recoveries, as well as the ICU utilization with the first infection on January 22 and the interventions beginning on March 22.

Using these adjusted parameters, how would the disease have evolved, if no interventions would have been imposed? Fig. 5 shows the dynamics under this assumption. As can be seen, the ICU utilization is highly exceeded (ca. 400K at its peak). Thus, this scenario would lead to a fairly higher number of deaths than shown in the figure because of significant overload of ICU capacities since we do not model maximum capacities but only reflect the required numbers. We now investigate the effect of these initial interventions between March 22 and April 19, without successive interventions, as illustrated in Fig. 6. We can see that the peak is only shifted to the right but not significantly reduced. This is a strong indication that the interventions cannot be released without substitution. Fig. 7 shows a scenario where the population is required to keep hygienic constraints (e.g., by using mouthpieces). It can be seen that the curve is clearly flattened, but the ICU utilization is still significantly too high (ca. 300K). The contact index shows the restrictions for the population by about 10% due to these hygienic measures but also that they can be reduced over time due to increasing immunization.

**Figure 5:**
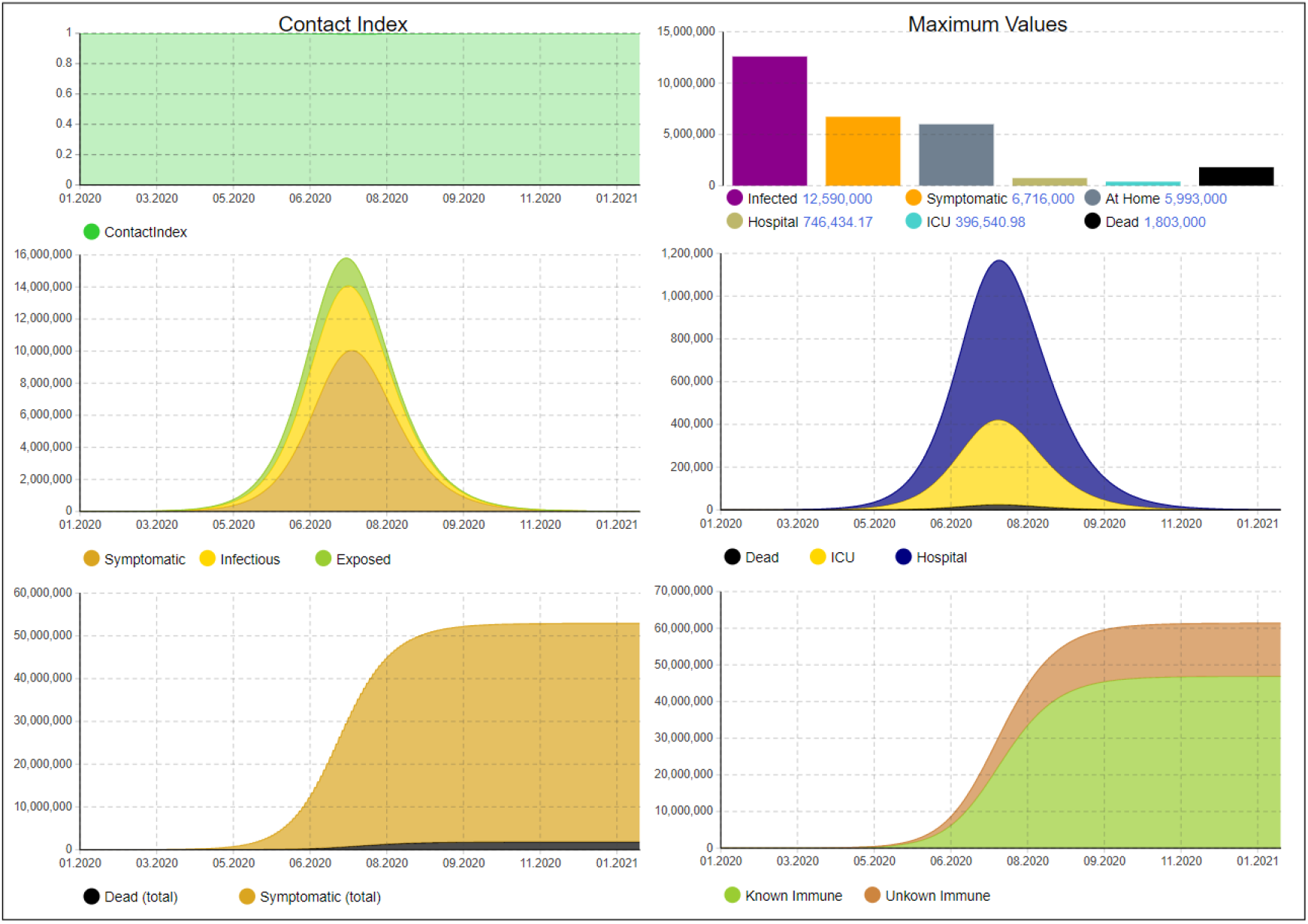
Disease dynamics without any intervention

**Figure 6:**
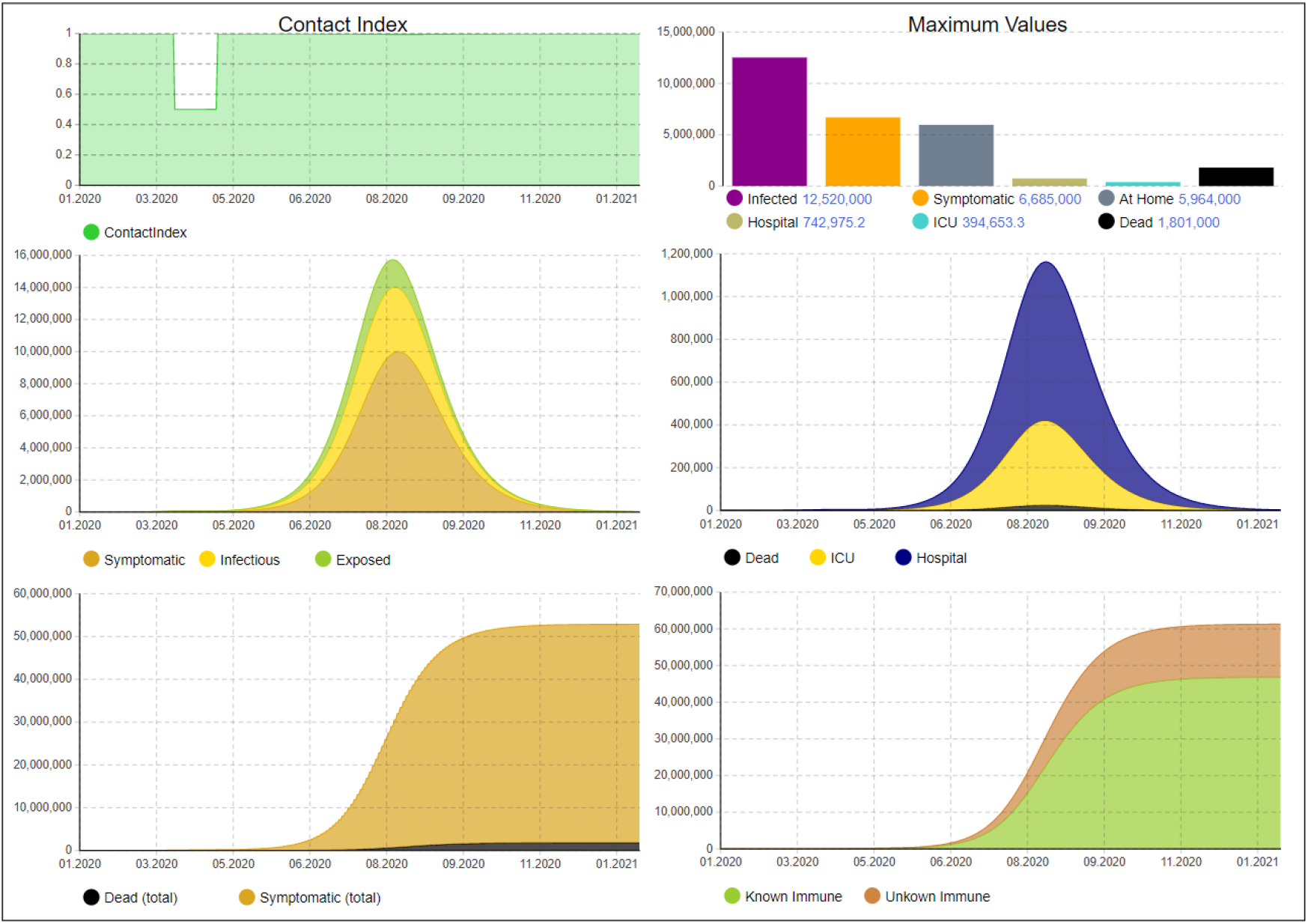
Disease dynamics without further interventions after April 19

**Figure 7:**
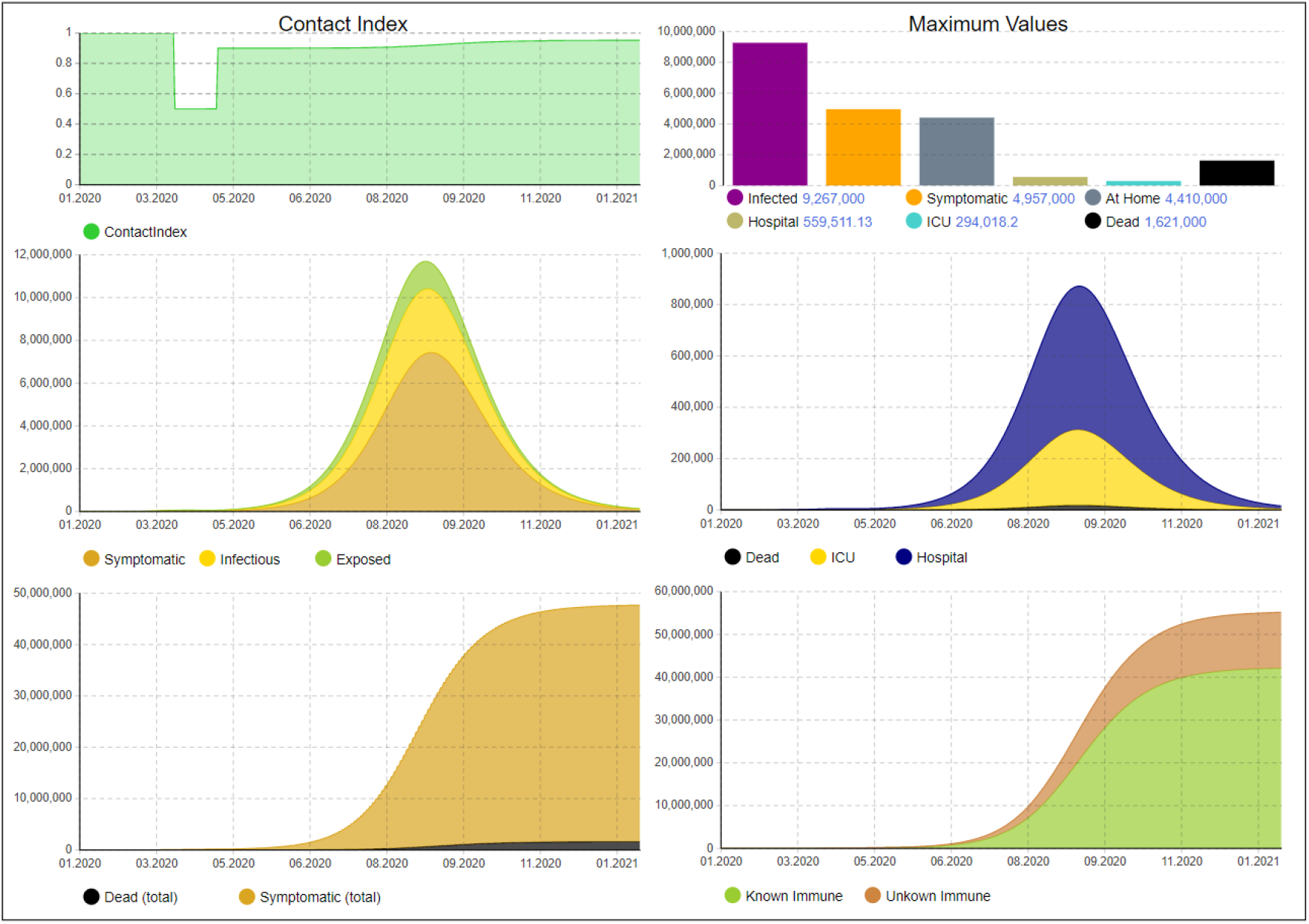
Disease dynamics with hygienic constraints but no other interventions after April 19

We use these insights to find an exit strategy which does not overload the health system, specifically ICU capacity. Contact reductions, as in the first lockdown, are triggered, if the current ICU occupation exceeds a certain threshold. Such additional adaptive restrictions periods should be short-term and can be released when the number falls below the threshold, however, in order to avoid oscillations, a minimum of two weeks for such reductions is assumed. This can easily be changed to other values or other indicators such as the number of deaths per day. This behavior can be directly transferred to reality by monitoring the utilization of the available ICU capacity and apply interventions if this number exceeds or falls below a certain threshold. Due to the delay when interventions affect the number of infections, we need to apply a threshold of 18.5K to ensure that the ICU capacity is not exceeded. The progression of the disease and the necessary interventions are illustrated in Fig. 8. As we can see, the model predicts that 9 additional intervals with contact restrictions would be necessary. The last intervention would end in March 2023 and also all hygienic measures could be dropped in 2023. Of course, the model can be adapted if new information about the disease is known, e.g., on seasonality. With an increasing number of recovered people after the infection, less people would be affected by interventions as reflected by the contact index. However, since this is a long period where interventions would be needed, we want to reduce the number of affected people as far as possible to reduce the economic and social impact but still introduce no additional risks for the population.

**Figure 8:**
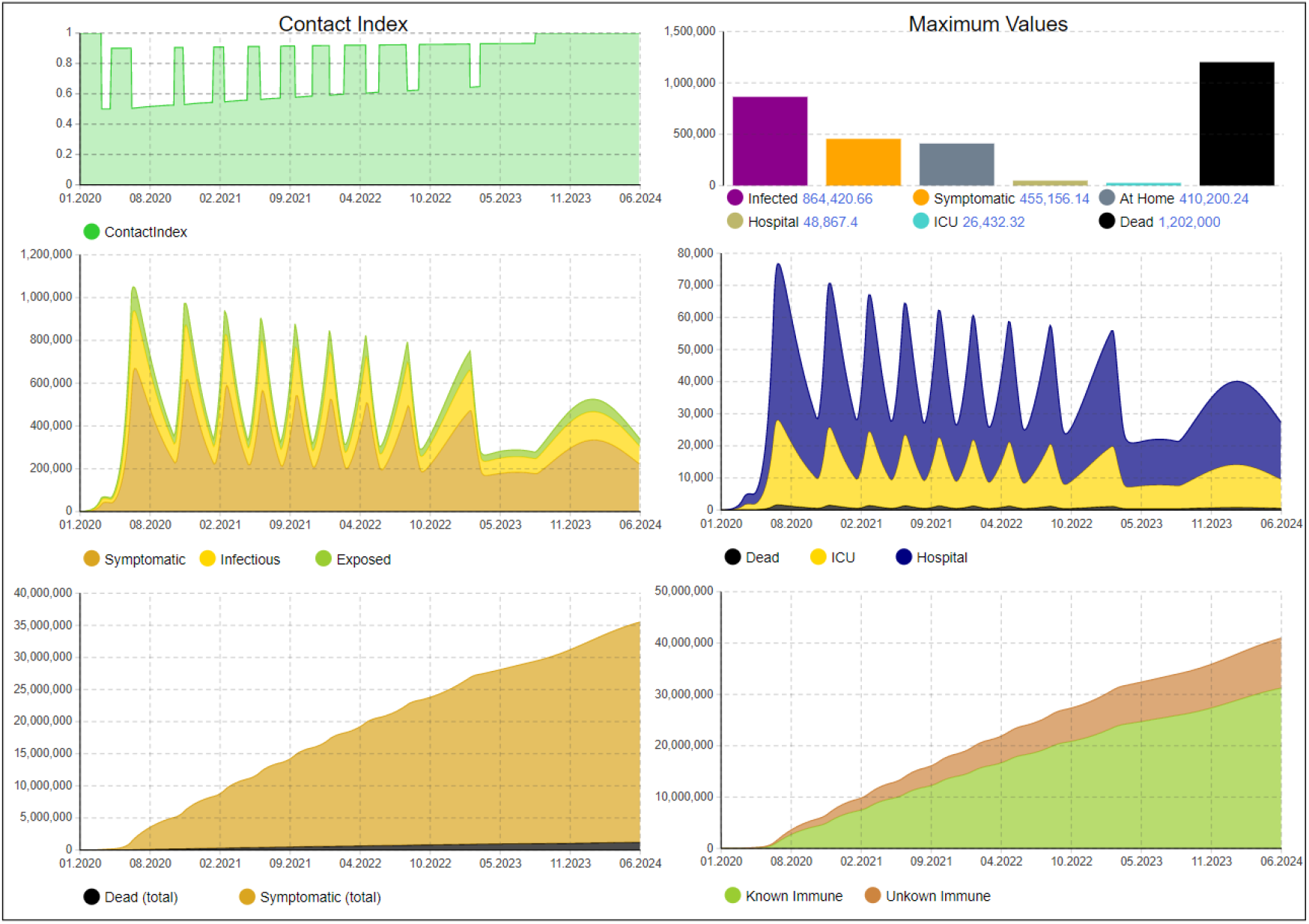
Disease dynamics with hygienic constraints and adaptive contact restrictions if ICU occupation exceeds a threshold

To achieve this, we now consider a scenario with AB tests, which we assume to start already after April 19. It is of course not yet realistic to have a significant capacity for AB tests in such a short term, but later in this year it could be accomplished if serious efforts are expended. We assume the same combination of interventions as in the last scenario (adaptive contact restrictions after the lockdown, hygienic measures). First of all, if test capacity would be unlimited, all unknown immunes would become known immunes, thus the orange stripe in Fig. 8 shows the maximum potential of AB tests which is 6.2M for this scenario. We now assume a test strategy in which preference is given to asymptomatic recovered persons and persons who had been symptomatic are not tested. Assuming that the asymptomatic recovered ones can be identified to some extend, we assign weights: *was_asymptomatic* → 40%, *was_symptomatic* → 0%, *asymptomatic* → 20%, *Prodromal* → 15%, *Exposed* → 15%, *Susceptible* → 10%.

Based on these assumptions the test capacity has been varied and the effects have been observed. It turns out that up to a capacity of 100K tests per day the number of people who can be excluded from contact reductions is increased significantly, afterwards less significant increase can be noticed. Even though requiring a high number of tests this does not seem to be completely out of scope if testing infrastructures can be scaled up. The effect with a capacity of 100K is illustrated in Fig. 9. As can be seen, up to 5.4M people can be excluded additionally from interventions with these tests (compared to 25.4M without tests). With a lower number of tests, e.g., 50K per day, we would still be able to exclude 4.4M people. If we apply these tests with preference to system-relevant groups, we can significantly improve public life (as a reference, ca. 1M nurses and related jobs are registered in Germany10). If we assume a more pessimistic value for the test sensitivity (75%), the effects are marginally reduced. With 50K tests per day, we would still be able to exclude 4M additional people. However, the effectiveness of AB testing significantly depends on the identification of asymptomatic recovered persons. If we assume that they cannot be identified (i.e., uniform weights for all stocks besides *was_symptomatic* which gets 0%), the number of additionally detected people with antibodies for 50K tests per day reduces to 2M. To achieve the similar effect as in Fig. 9 before, the number of tests need to be increased even up to 250K. This shows that tracking might be a valuable tool for future exit strategies.

**Figure 9:**
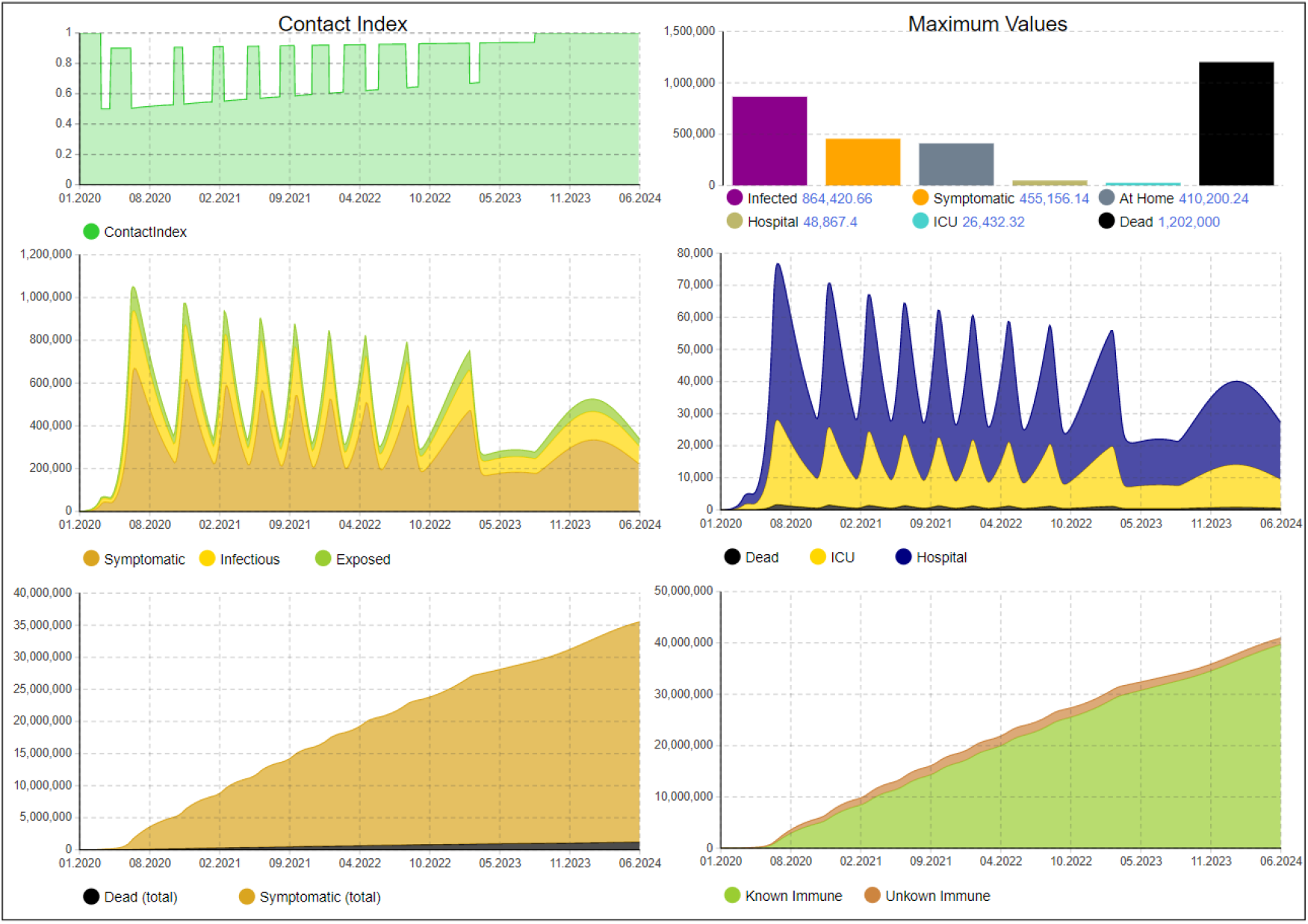
Disease dynamics with hygienic constraints and adaptive contact restrictions if ICU occupation exceeds a threshold and application of antibody tests

The importance of AB tests would be even greater if the disease spreads more drastically, e.g., due to non-optimal interventions. This in turn would lead to more infections and in turn to more asymptomatic recovered and thus increase the likelihood of finding people with antibodies, making the tests more effective and excluding more people from interventions. However, we have not added figures for these scenarios because the ICU capacities would be exceeded and the number of deaths would increase significantly, as discussed in the previous scenarios.

Fig. 10 and 11 demonstrate how the virus spreading would be affected if a seasonality factor is in place, meaning that the virus spreads less in summer and more in winter months, or if a significant portion of the population is immune to the virus. The seasonality is modeled as sine curve, as also explained in the Appendix. The figures show the dynamics with no antibody testing in place, the number of immune people and thus the contact index could be improved with the previously described measure by the number of unknown immune people tracked by the bottom right curve. We omitted the graphics for space reasons but the number of additionally identified immune people behaves nearly identically as with the effect shown in Fig. 8 and 9. As we can see, the dynamics of the virus are similar, the disease will still spread fast after April 19 and would exceed ICU capacities. The main difference is the number of interventions that will be required, which reduces from 9 to 6 (for seasonality) or 5 (for initially immune people). If an initial immunity of one third of the population is assumed, the last intervention is already in November 2021. To optimally propose interventions, our model can thus be adapted when more information for this behavior is known. However, currently, we assume the worst-case behavior without seasonality and no initial immunization.

**Figure 10:**
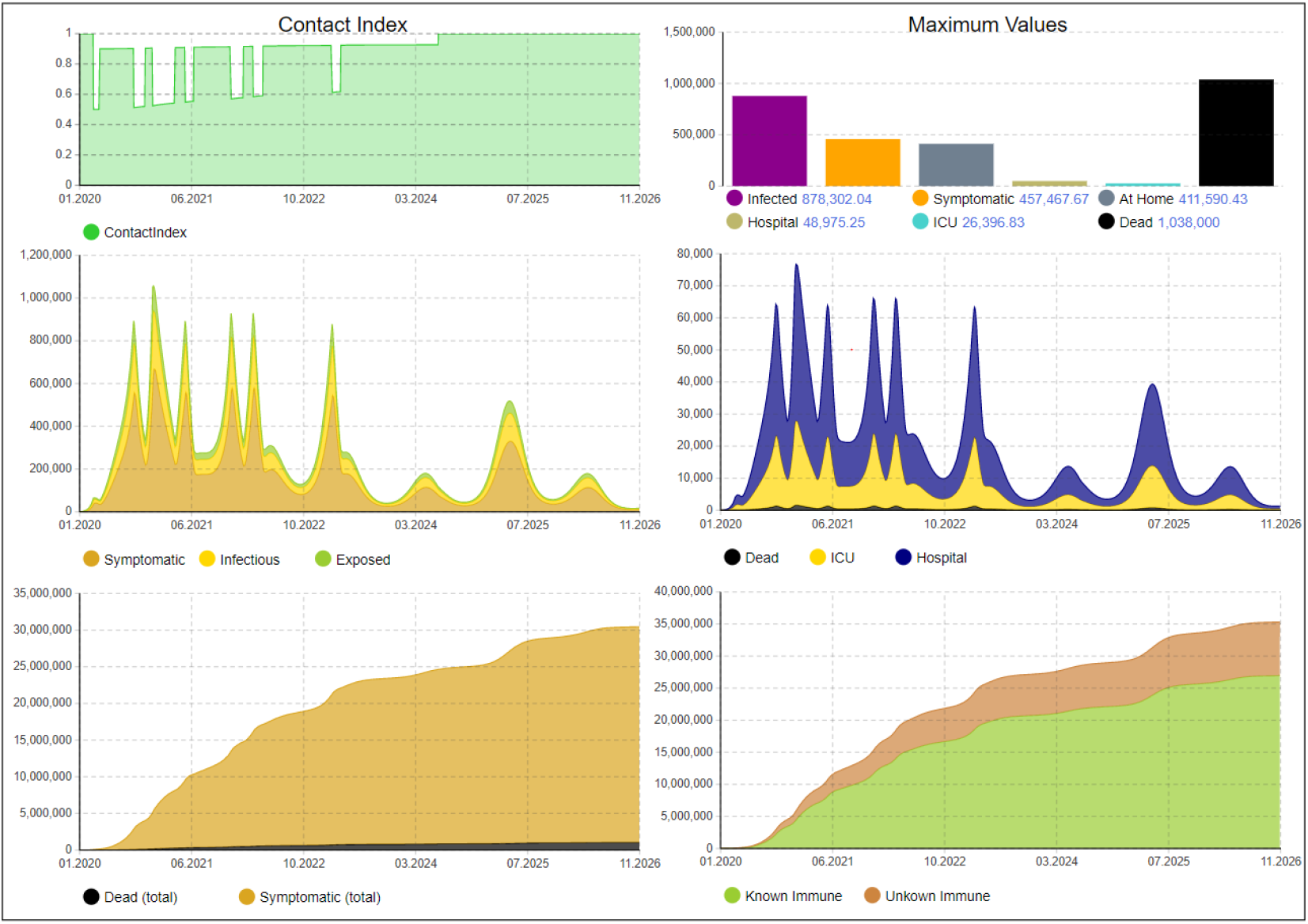
Disease dynamics if the virus is more infectious during winter months, less in summer (*R*_0_ fluctuates between 3.1 and 2.1 with sine curve)

**Figure 11:**
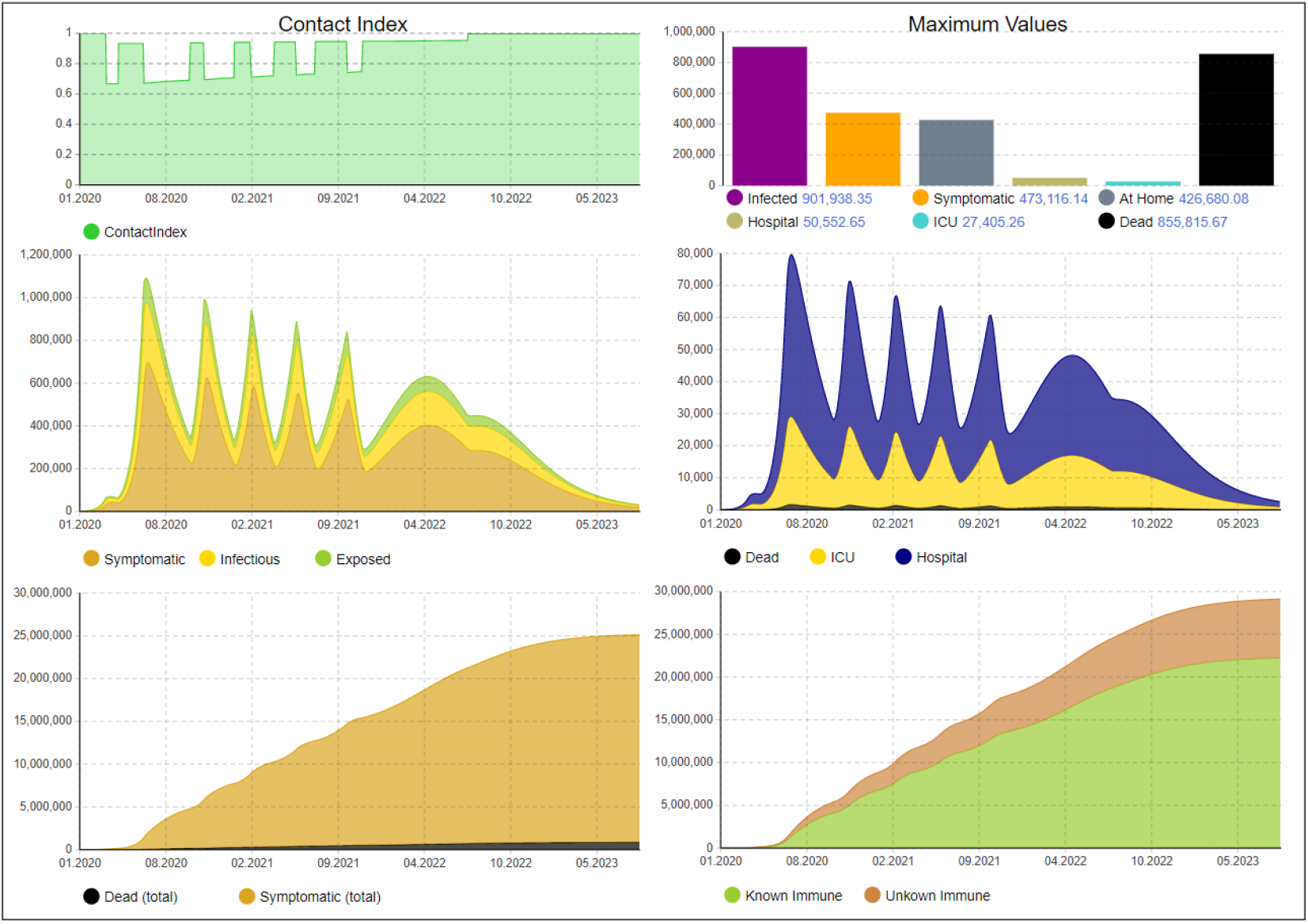
Disease dynamics if one third of the population is generally immune to the virus

## 7 Conclusions

We have presented a system dynamics (SD) and an agent-based simulation (ABS) model for COVID-19. The models can represent the disease dynamics on different abstraction levels and can also be used to study governmental interventions and the effects of antibody tests. Starting from parameters suggested by other COVID-19 models recently presented we adapted the parameters to published data for Germany and investigated derived scenarios from that. Based on these assumptions about parameters and the SD model, which is of course a simplified abstraction of reality, we could gain the following key insights for a country like Germany:

- Without enacting interventions such as the lockdown in March 2020, the disease would cause significant overload of the health system with many deaths expected. This is in agreement with the findings of other COVID-19 models.
- If the lockdown would just be released after 4 weeks in April and a return takes place to the status before lockdown, the disease can be expected to rebound just with a slight delay corresponding to the time of the lockdown, meaning that a little time has been gained in order to prepare the health system better but with the same potentially large number of deaths.
- If hygienic measures are put in place after lockdown, a slight mitigation can be expected but this alone would not be sufficient to defeat the disease.
- This however can be achieved with repetitive short term contact reductions similar to the current lockdown; such reductions can be triggered adaptively, if relevant figures (such as death rates, need for ICU, etc.) exceed a threshold. With additional hygienic measures the situation can be enhanced further. However, we can expect that this situation (repetitive short term lockdowns and hygienic measures) needs to be in place for the next two or three years until herd immunity can be obtained (if vaccination is not available before).
- The effects of antibody tests would add significant benefit in order to exclude people with antibodies from the contact reductions. Results show that already a moderate infrastructure for antibody tests (50K per day for Germany) would lead to significant improvements bringing ca. 4.4M people back to public life compared to scenarios without such possibilities. With a higher test capacity (e.g., 100K) this number could be pushed to more than 5.4M. Digital tracing could improve the efficiency of AB tests. If immunity will be lost after some time, AB testing would become even more important.
- Seasonality of the disease and general immunization would lead to significant mitigation effects while still requiring the described mechanisms.
- A contact index to condense the social contact abilities for all people has been defined, it can illustrate well the effect of the combination of all measures.

Both models can be extended in several ways in order to study the interaction between further effects of the disease and more sophisticated exit strategies, such as benefits from PCR tests as well, age- and risk-dependent severity and fatality rates, more detailed consideration of infectiousness during the phases of illness, influence of viral load on severity, differentiation of measures for vulnerable groups of people, etc. One promising approach for this is to combine both models to a hybrid simulation model as in [5] in which both general trends and details for regions, different groups of persons, and more exit strategies can be studied. The models can be adapted to latest data as the pandemic is progressing in order to adapt interventions accordingly. The integration of digital tracing for epidemic control (alerting people in case of previous contacts with people who have been infected) [2] into the set of measures is also promising and can be investigated with the presented models.

## Data Availability

All data is given in the paper directly

## A SD model details

We provide all initial stock values, equations, and parameters of the SD model.

### A.1 Initial stock values

All stocks besides the following are initially set to zero:

- *S* = (1 *− p*_*init*_*immune*_) *∗ N − IF*
- *P* = *IF*
- *Ak* = *p*_*init*_*immune*_ *∗ N*

### A.2 Differential equations of the output flows

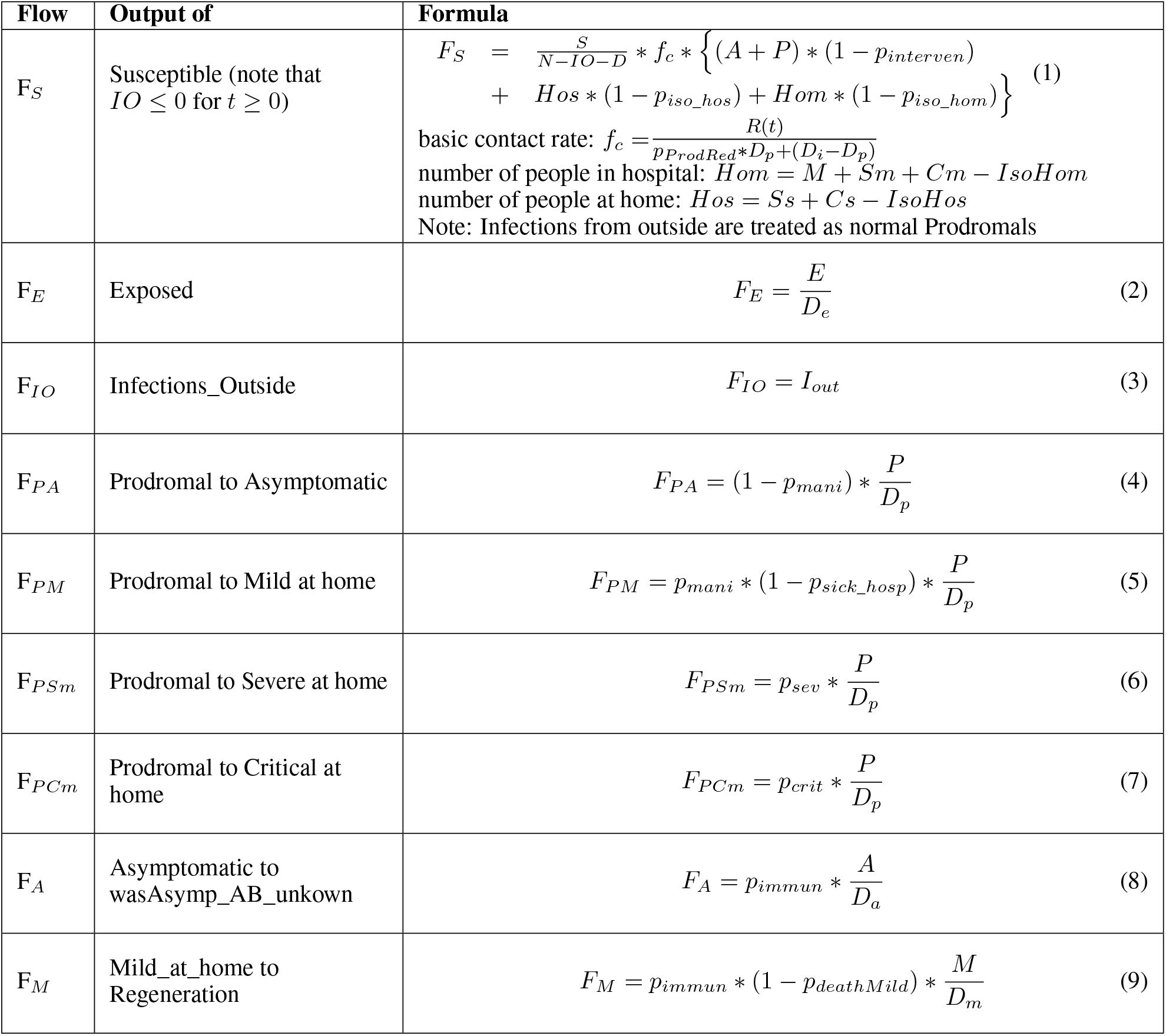
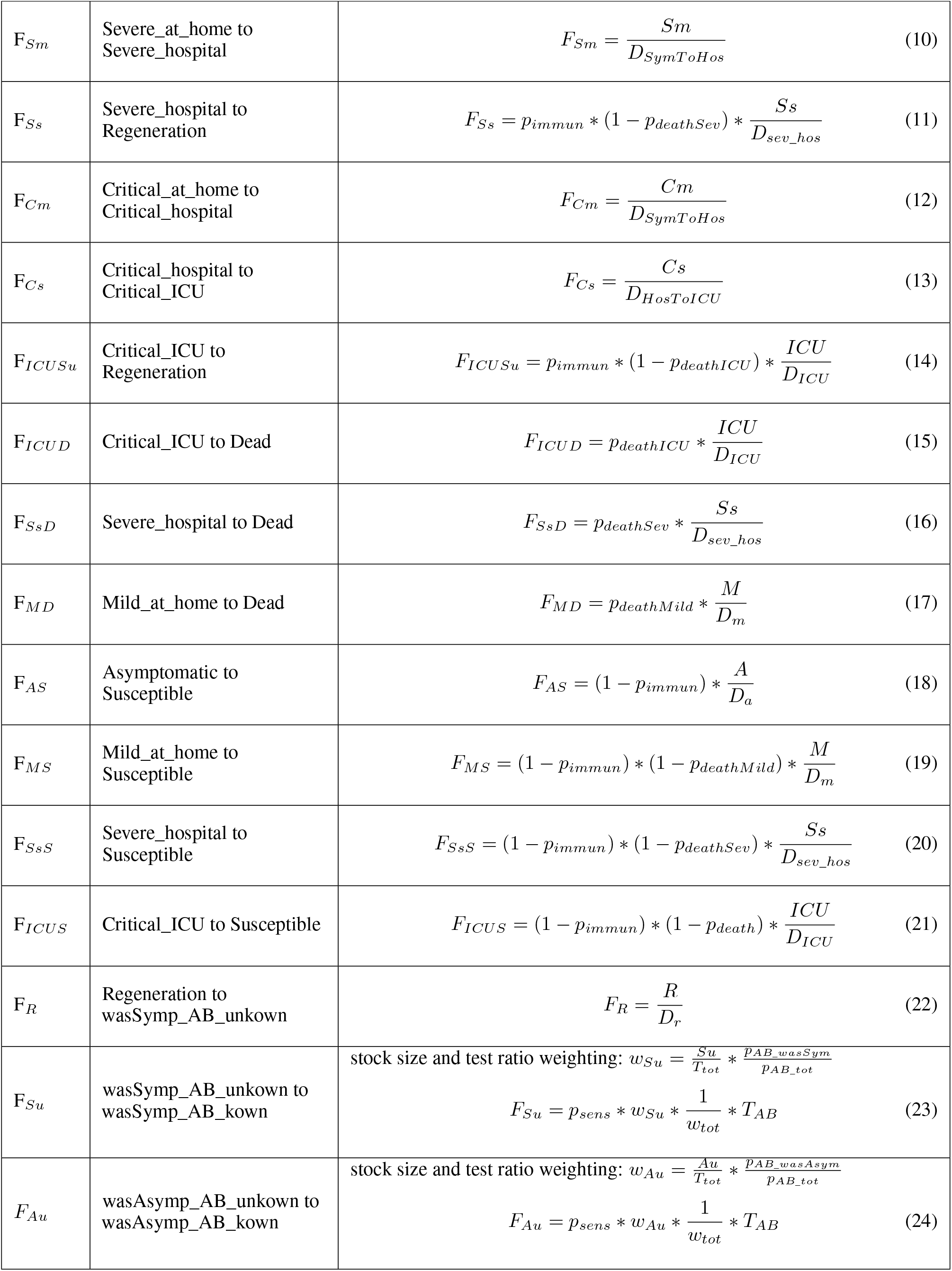

### A.3 Parameters

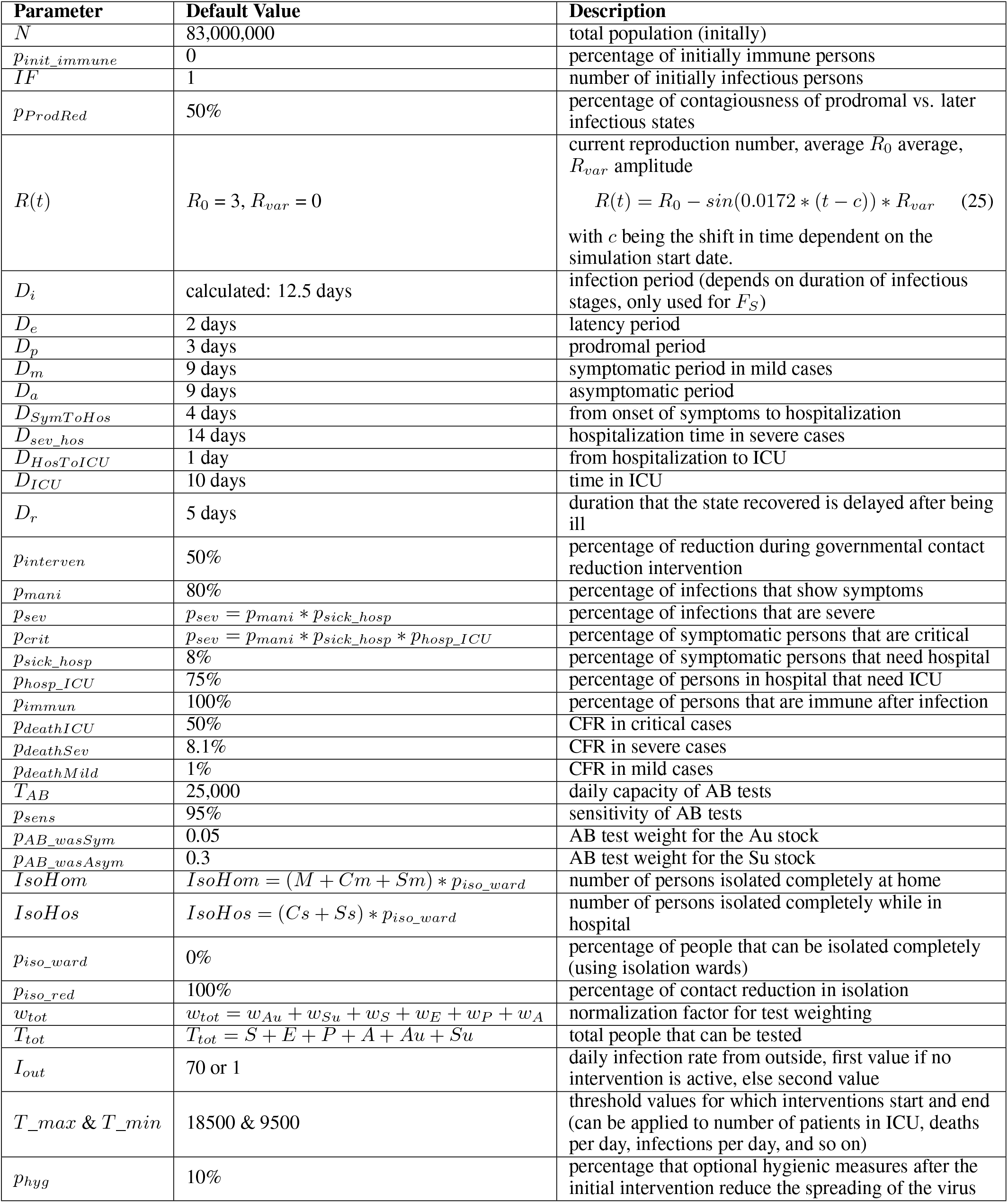

## Footnotes

1 https://coronavirus.jhu.edu/map.html

2 https://www.anylogic.com/

3 covidsim.eu

4 http://www.dwh.at/de/neues/wie-man-die-epidemie-berechnen-kann/

5 https://exchange.iseesystems.com/public/isee/covid-19-simulator/

6 https://forio.com/app/jeroen_struben/corona-virus-covid19-seir-simulator/

7 https://www.rki.de/DE/Content/InfAZ/N/Neuartiges_Coronavirus/Steckbrief.html

8 https://www.intensivregister.de/#/intensivregister

9 https://coronavirus.jhu.edu/map.html

10 https://www.bundesgesundheitsministerium.de/themen/pflege/pflegekraefte/beschaeftigte.html

